# Gargle-Direct: Extraction-Free Detection of SARS-CoV-2 using Real-time PCR (RT-qPCR) of Saline Gargle Rinse Samples

**DOI:** 10.1101/2020.10.09.20203430

**Authors:** Vijay J. Gadkar, David M. Goldfarb, Virginia Young, Nicole Watson, Linda Hoang, Tracy Lee, Natalie Prystajecky, Ghada N. Al-Rawahi, Jocelyn A Srigley, Peter Tilley

## Abstract

**Background:** Saline mouth rinse/gargle samples have recently been shown to be a suitable option for swab-independent self-collection for SARS-CoV-2 diagnosis. We sought to evaluate a simplified process for direct reverse transcriptase PCR (RT-qPCR) testing of this novel sample type and to compare performance with routine RT-qPCR using automated nucleic acid extraction.

**Methods:** Clinical saline mouth rinse/gargle samples were subjected to automated nucleic acid extraction (“standard method”), followed by RT-qPCR using three assays including the FDA authorized US-CDC’s N1/N2 assay, which was the reference standard for determining sensitivity/specificity. For extraction-free workflow, an aliquot of each gargle sample underwent viral heat inactivation at 65 °C for 20 minutes followed by RT-qPCR testing, without an intermediate extraction step. An in-house validated RT-qPCR lab developed test (LDT), targeting the SARS-CoV-2’s S/ORF8 genes (SORP triplex assay) and the N1/N2 US-CDC assay was used to evaluate the extraction-free protocol. To improve the analytical sensitivity, we developed a single-tube hemi-nested (STHN) version of the SORP triplex assay.

**Results:** A total of 38 SARS-CoV-2 positive and 75 negative saline mouth rinse/gargle samples were included in this evaluation. A 100% concordance in detection rate was obtained between the standard method and the extraction-free approach for the SORP assay. An average increase of +2.63 to +5.74 of the cycle threshold (C_T_) values was observed for both the SORP and N1/N2 assay when extraction-free was compared between the standard method. The average ΔC_T_ [ΔC_T_=C_T(Direct PCR)_-C_T(Extracted RNA)_], for each of the gene targets were: S (ΔC_T_= +4.24), ORF8 (ΔC_T_=+2.63), N1 (ΔC_T_=+2.74) and N2 (ΔC_T_=+5.74). The ΔC_T_ for the STHN SORP assay was +1.51 and −2.05 for the S and ORF8 targets respectively, when extracted method was compared to the standard method.

**Conclusion:** Our Gargle-Direct SARS-CoV-2 method is operationally simple, minimizes pre-analytical sample processing and is potentially implementable by most molecular diagnostic laboratories. The empirical demonstration of single-tube hemi-nested RT-qPCR, to specifically address and alleviate the widely-acknowledged problem of reduced analytical sensitivity of detection of extraction-free templates, should help diagnostic laboratories in choosing Gargle-Direct protocol for high-throughput testing.

## INTRODUCTION

Currently, the widely used diagnostic method for detection of SARS-CoV-2 is based on swabbing the patient’s nasopharynx using a flocked swab. Post-swabbing, transport of the nasopharyngeal flocked swab (NPFS) material in a tube containing viral transport medium (VTM), followed by RNA isolation/purification and subsequent analysis by reverse transcriptase polymerase chain reaction (RT-qPCR) is currently the most common method of clinical detection of SARS-CoV-2 virus (Fig. 1A). This procedure, however, has proven to be difficult in mass-scale testing for SARS-CoV-2, primarily due to its resource intensive nature, as it requires a health care worker (HCW) wearing a personal protective equipment (PPE) to collect the sample and its associated discomfort to the patient while performing the collection. Moreover, unexpected challenges such as shortage of NPFS collection devices in many jurisdictions or sub-standard quality of the device itself, have resulted in difficulties in implementing mass scale testing of SARS-CoV-2. To address this, alternative sample types, preferably those which can be self-collected have been proposed. Saliva has recently been shown to perform similarly to the NPFS gold standard (Azzi et al., 2020; Ranoa et al., 2020; Vogel et al., 2020; Wyllie., 2020). In addition to being self-collectible, saliva samples have also been used in a directly in the RT-qPCR reaction, without recourse to a discrete RNA extraction procedure (Vogel., 2020) This feature is extremely appealing since large scale RNA extraction is not only cumbersome and expensive, but recent instances of supply chain bottlenecks in the RNA extraction reagents (Esbin et al., 2020), have made it difficult for clinical laboratories to implement large scale SARS-CoV-2 testing (Bruce et al., 2020; Fomsgaard and Rosenstierne, 2020; Merindol et al., 2020).

**Fig. 1.**
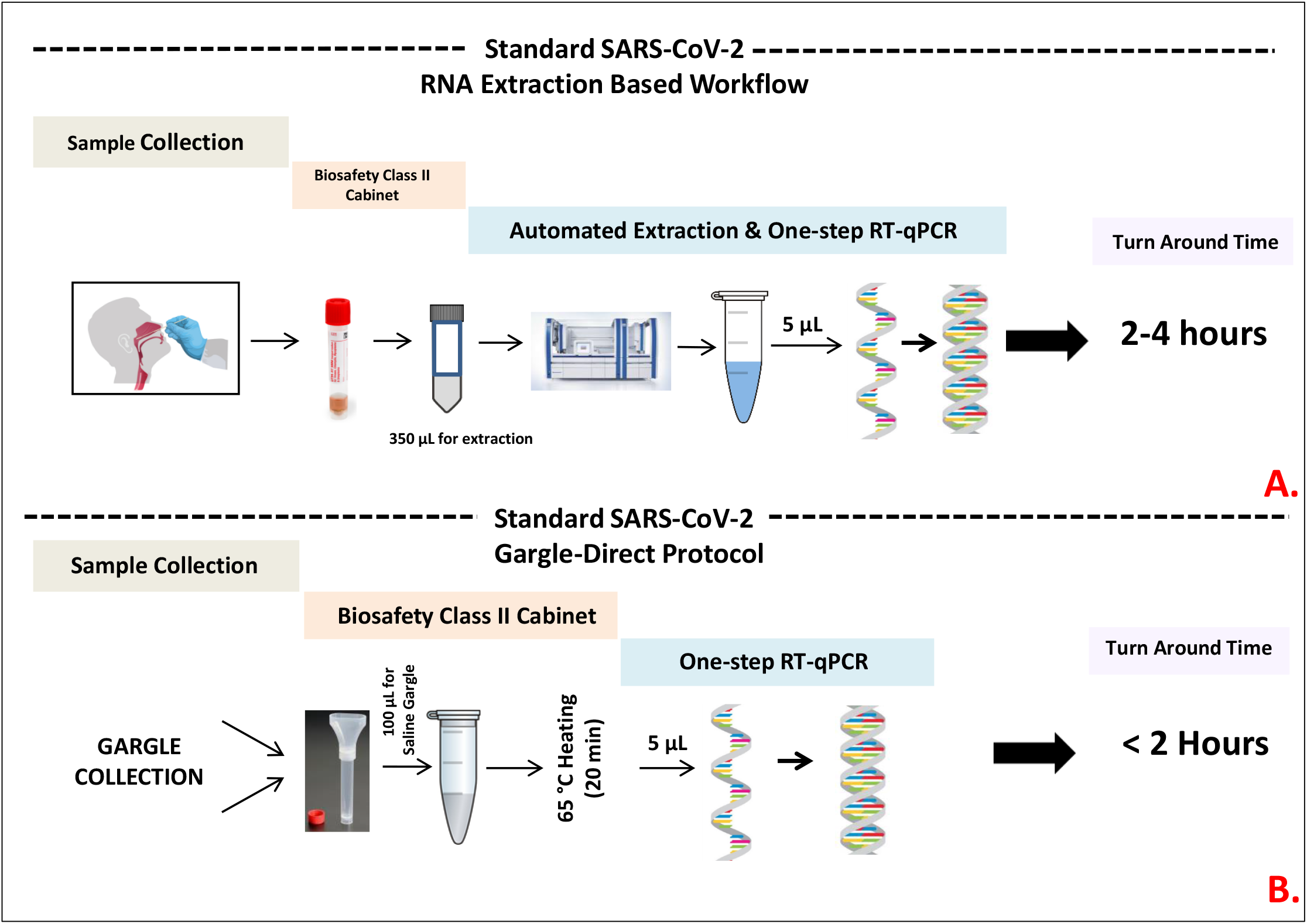
Diagram describing the standard RNA extraction-based protocol (A.) and extraction-free protocol. The turn around time (TAT) is an estimated time taken to process in parallel, a single batch of 24 samples by the standard and extraction-free approach.

Using saliva as a sample type in standard RNA extraction (manual/automated process) based or direct (extraction-free) RT-qPCR protocols has however, proven to be challenging, owing to saliva’s heterogeneity and viscosity. Steps to reduce sample viscosity, such as use of liquefaction buffers containing mucolytic agent for e.g. dithioerythritol (Hammerschlag et al., 1980), have been recommended, in order to make it amenable for dispensing (both manual and automated) for downstream processes. In direct saliva-to-RT-qPCR protocols, some of the validated pre-analytical steps to reduce the viscosity and inactivate the virus include, 1:1 dilution of saliva in either TE/TBE buffer, followed by heating at 95 °C for 30 minutes (Ranoa et al., 2020), or incubation of the saliva sample with a deproteinizing agent (Proteinase-K), followed by heat inactivation at 95 °C (Vogel et al., 2020). These pre-analytical steps can be challenging for high-throughput laboratories to implement.

We recently showed that self-collected saline mouth rinse/gargle samples had similar performance as NPFS for detection of SARS-CoV-2 in adults and children presenting with outpatient illness (Goldfarb et al., 2020). In the present work, we seek to evaluate this sample type with a simplified process for detecting SARS-CoV-2, without RNA extraction (extraction-free PCR; Fig. 1B). Performing direct gargle-to-RT-qPCR, henceforth referred to as “Gargle-Direct”, requires only a simple heating step at 65 °C for 20 minutes to inactivate the virus (Wang et al., 2020). This heating step was incorporated primarily for biosafety reasons to enable automated processing (e.g. on open liquid handler platforms) while avoiding high temperatures which are harder to automate and can negatively impact RNA stability (Zou et al., 2020; Pastorino et al., 2020). To compensate for the expected decrease in efficiency of the SARS-CoV-2 RT-qPCR assay due to the use of unpurified lysed templates, we also explored the use of single-tube hemi-nested real-time-qPCR (STHN-RT-qPCR) to enhance the overall sensitivity of our SARS-CoV-2 detection assay.

## MATERIALS & METHODS

### Clinical specimens & Nucleic acid extraction

Gargle specimens submitted for routine SARS-CoV-2 testing, at the Microbiology & Virology Laboratory of BC Children’s Hospital were used in the study. The standard method for detection of SARS-CoV-2 was as follows: extraction of total nucleic acid (TNA) from 200 µL of sample on the QIAsymphony (Qiagen, Germantown, MD, USA) automated extraction platform using the DSP Virus/Pathogen kit (Qiagen). The eluate (ca. 80 µL) was submitted for RT-qPCR assay. Post-testing, the residual gargle samples were anonymized and used for developing the Gargle-Direct protocol. Ninety-four samples used for this evaluation had already been stored at −80 °C and needed to be thawed for testing. The remainder were tested prospectively without having been subjected to a freeze thaw cycle. Approval was obtained from the BC Children’s and Women’s Hospital Research Ethics Board for this study (H20-02538).

### Extraction-free PCR

Aliquots of the anonymized gargle samples (100 µL) were heat treated at 65 °C for 20 minutes to inactivate the virus. Post-heating, the sample was allowed to cool at room temperature for 5 minutes and 5 µL of the sample was directly added to the RT-qPCR reaction.

### SARS-CoV-2 RT-qPCR assays

All of the RT-qPCR assays were performed on the ABI Fast 7500 real-time PCR system (Life Technologies, Carlsbad, CA) machines. The total reaction volume was 20 µL, including 5 µL of the template. For the standard RNA extraction-based approach, the 4X TaqMan Fast Virus 1-step Master Mix (ThermoFisher: cat No 4444434) was used across all the different assays. The cycling condition of 50 °C for 5 minutes (Reverse Transcription), 95 °C × 20 seconds (Enzyme Activation) followed by 45 cycles of 95 °C at 3 seconds and 60 °C at 30 seconds, as recommended by the manufacturer was used. For the extraction-free approach, the Luna™ One-Step Universal RT-qPCR master mix (New England Biolabs, Whitby, ON; Cat No: E3006E) was used. The following cycling parameters, as recommended by this manufacturer were used: 55 °C for 10 minutes (Reverse Transcription), 95 °C × 1 min (Enzyme Activation) followed by 45 cycles of 95 °C at 10 seconds and 60 °C at 30 seconds. For STHN-RT-qPCR, high-temperature (70°C) primary cycling (15 cycles) was incorporated before the standard 45 cycle amplification. ΔC_T_ in the present study was defined as the difference between the C_T_ values obtained between Direct PCR and Extracted methods [ΔC_T_=C_T(Direct PCR)_-C_T(Extracted RNA)_].

### N1/N2/RNP US-CDC’s Assay

The US CDC’s N1 (2019-nCoV_N1) and N2 (2019-nCoV_N2)/RNase P (RNP) primer-probe sets (Table. 1) were used as the reference standard. The interpretation of N1/N2/RNase P results was done as described as per US-CDC’s guidelines (US-FDA 2020). For statistical analysis, any negative gene target value was assigned a C_T_ value of 41.

**Table. 1.**
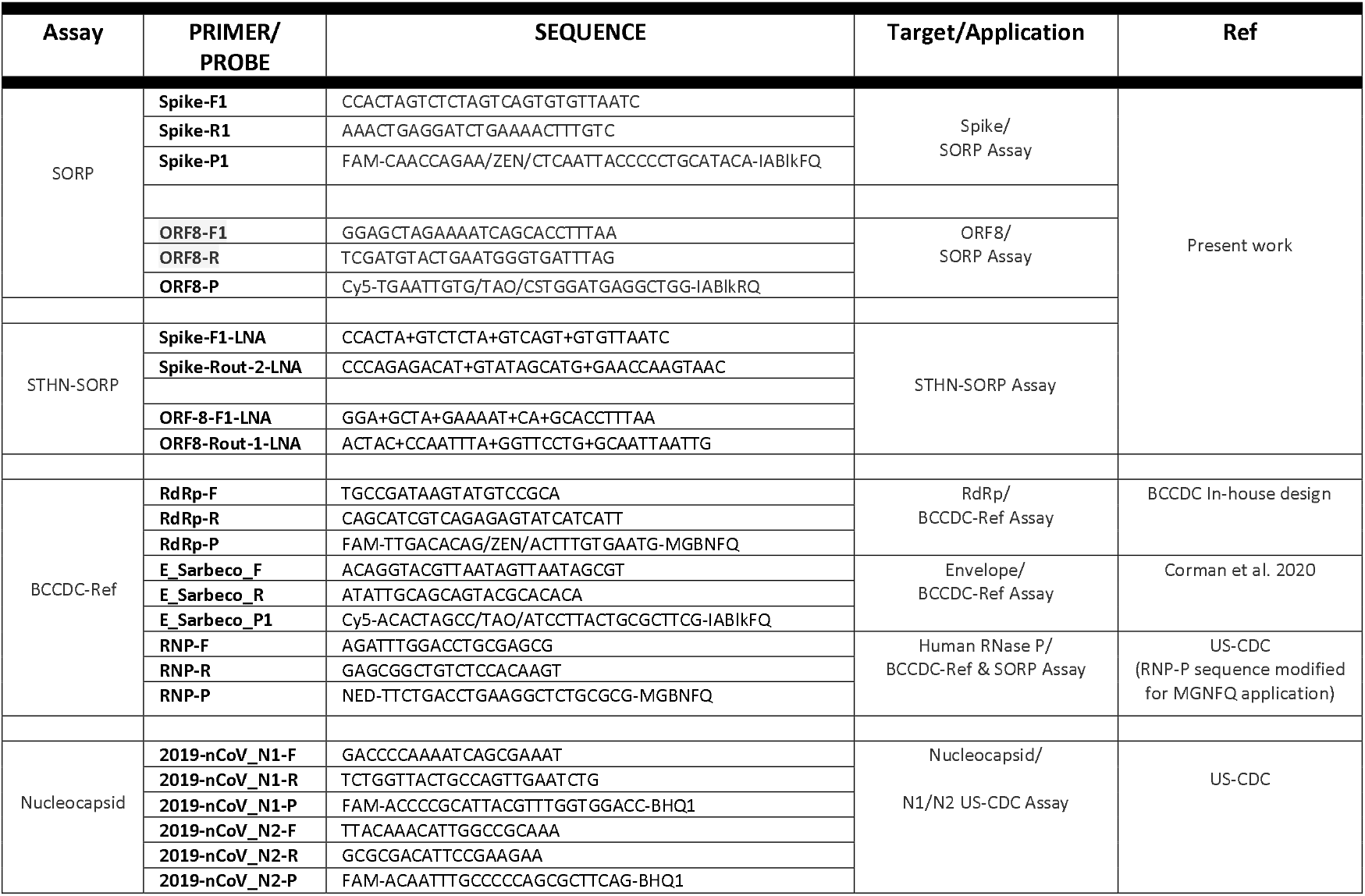
List of RT-qPCR primer and TaqMan™ probes used in the present study. The “+” sign denotes a locked nucleic acid (LNA) base. STHN: Single tube hemi-nested

### BCCDC-Ref Assay

The British Columbia Centre for Disease Control’s (BCCDC’s) SARS-CoV-2 triplex assay, henceforth referred to as “BCCDC-Ref,” was implemented as described (LeBlanc et al., 2020). This is a multiplex assay and targets the SARS-CoV-2’s RNA dependent RNA polymerase (RdRp), Envelope *(E)* genes, with the human RNase P as the internal control (Table. 1). For assay interpretation, single gene targets positives were interpreted as indeterminate and for sensitivity/specificity calculation as positive. For statistical analysis, any negative gene target value was assigned a C_T_ value of 41.

### SORP (Spike, ORF8, RNAse P) Assay

The SORP triplex assay (<u>S</u>pike, <u>OR</u>F8, and human RNase <u>P</u>), an in-house assay, validated against both the reference US-CDC Nucleocapsid assay and the BCCDC-Ref assay was also used (Table 1). This multiplex assay targets the SARS-CoV-2’s Spike (S) and ORF8 genes with the human RNase P as the internal control (manuscript in preparation). The SORP RT-qPCR assay reaction consisted of (20 µL) with the following concentration of the primer and probes: Spike-F1/R1 (0.3 µM), Spike-P1 (0.2 µM), ORF8-F1/ORF8-R (0.4 µM), ORF8-P (0.2 µM), RNase P-F/R (0.05 µM) and RNase P-P (0.15 µM). The primers and probes are listed in Table 1.

A single-tube hemi-nested version of the SORP-RT-qPCR assay, henceforth referred to as “STHN-RT-qPCR” SORP assay, was also used. This assay consisted of the same reaction components used for standard the SORP assay, except for two changes (a) addition of four locked nucleic acids (LNA) modified primer for increased Tm, for the primer to anneal at 70 °C and (b) a “two-stage” PCR cycling condition. These additional primers are listed in Table 1. The concentration of these additional LNA hemi-nested primers were: Spike-F1-LNA/Spike-Rout-2-LNA (0.05 µM) and ORF8-F1-LNA/ORF8-Rout-1-LNA (0.05 µM). The hemi-nested PCR cycling conditions consisted of: 55 °C for 5 minutes (RT), 95 °C × 1 min (Activation), 15 cycles of 95 °C for 10 sec and 70 °C for 60 seconds (Primary cycling), followed by 45 cycles of 95 °C for 3 seconds and 60 °C for 30 seconds (Secondary cycling). For data interpretation purpose, single gene targets for both standard SORP and STHN-RT-qPCR assays, were interpreted as indeterminate and for sensitivity/specificity calculation as positive. For statistical analysis, any negative gene target value was assigned a C_T_ value of 41.

## RESULTS

A total of 38 SARS-CoV-2 positive and 75 SARS-CoV-2 negative gargle samples previously tested by the BCCDC-Ref assay were included in this study. Samples were collected between 2^nd^ August 2020 and 26^th^ September 2020. Fourteen of the positive samples were prospectively collected without going through a freeze thaw cycle prior to experimental testing.

### Diagnostic performance of N1/N2 & SORP Triplex assay

All 38 positive and 75 negatives gargle samples tested clinically by the BCCDC-Ref assay showed 100% concordance on the N1/N2 US-CDC assay using the standard extracted RNA method. The average cycle threshold (C_T_) values of the 38 positive gargle samples on the N1/N2 assay were: N1=27.15 and N2=27.61. The human RNase P was detected in all the 38 positives and 75 negative samples, indicating adequate sample collection.

The N1/N2 US-CDC assay, detected with both the N1 and N2 targets in the 38 positive saline mouth rinse/gargle samples using the Gargle-Direct approach however, four “invalid results” (N1<40 and N2 >40) were recorded (sample no: FS5, FRS10 and FRS16, FRS18) (Table. 2). The average C_T_ value of the 38 positive gargle samples detected by the standard RNA extraction-based protocol were: N1=27.15 and N2=27.61. (Table. 2). For the same cohort of positive gargle samples tested on the Gargle-Direct approach, the average C_T_ values for the N1 and N2 gene targets were: 29.89 and 33.35 respectively (Table. 3). This represented a ΔC_T_ of +2.74 and +5.74, for the N1 and N2 gene targets respectively (Table. 3) which was statistically significant using two-sided paired-sample t test (P<0.0001; Fig. 2). The RNase P internal control was detected in all the positive and negative gargle samples, in both standard and extraction-free methods. Overall, the performance of the N1/N2 US-CDC assay on the Gargle-Direct protocol was: sensitivity of 100% (95% CI: 89.72% to 100.0%), specificity of 100% (95%CI: 95.20% to 100.00%), Accuracy: 100% (95%CI: 96.67% to 100.0%).

**Table. 2.** C_T_ values obtained for each of the gene targets for the BCCDC-Ref, SORP (standard and single-tube Hemi-nested) and the N1/N2 assay. Standard method: Extracted RNA method, Gargle-Direct: Extraction free method, C_T_ value of 41 = no signal detected. Prospective*: Samples did not undergo a freeze-thaw cycle.

|  |  |  | Standard Method |  | Standard Method |  | Gargle-Direct |  | Gargle-Direct |  | Standard Method |  | Gargle-Direct |  |  |
| --- | --- | --- | --- | --- | --- | --- | --- | --- | --- | --- | --- | --- | --- | --- | --- |
|  |  |  | BCCDC-Ref |  | SORP |  | SORP |  | SORP (Hemi-Nested) |  | N1/N2-US CDC |  | N1/N2-US CDC |  |  |
|  |  | TYPE | RdRp | E gene | S | ORF8 | S | ORF8 | S | ORF8 | N1 | N2 | N1 | N2 | RNAse P |
| 1 | FS1 | Frozen | 31.17 | 31.56 | 30.82 | 32.68 | 34.51 | 34.72 | 32.21 | 29.90 | 29.91 | 30.53 | 34.35 | 37.66 | 29.35 |
| 2 | FS2 | Frozen | 22.30 | 21.88 | 21.09 | 23.08 | 25.93 | 25.80 | 22.85 | 21.15 | 20.56 | 20.95 | 31.51 | 26.20 | 26.56 |
| 3 | FS3 | Frozen | 28.28 | 28.49 | 27.73 | 29.99 | 31.17 | 33.01 | 28.70 | 27.69 | 27.62 | 27.77 | 30.99 | 33.35 | 29.00 |
| 4 | FS4 | Frozen | 26.60 | 26.60 | 26.09 | 28.31 | 29.22 | 30.09 | 27.01 | 25.79 | 25.41 | 26.13 | 27.29 | 31.60 | 29.14 |
| 5 | FS5 | Frozen | 24.67 | 24.89 | 23.80 | 29.53 | 28.19 | 31.45 | 24.26 | 25.02 | 23.30 | 24.89 | 26.37 | 41.0 | 28.36 |
| 6 | FS6 | Frozen | 24.80 | 25.10 | 24.40 | 26.28 | 29.19 | 29.68 | 27.66 | 27.90 | 23.74 | 23.89 | 26.97 | 29.45 | 26.18 |
| 7 | FS7 | Frozen | 26.90 | 27.21 | 25.87 | 26.30 | 30.46 | 30.77 | 28.01 | 25.97 | 25.20 | 26.96 | 28.10 | 30.80 | 28.61 |
| 8 | FS8 | Frozen | 30.60 | 30.80 | 31.20 | 30.12 | 32.07 | 33.96 | 30.41 | 28.76 | 29.43 | 30.23 | 31.83 | 34.14 | 29.89 |
| 9 | FS9 | Frozen | 28.98 | 29.38 | 27.89 | 28.45 | 30.46 | 32.26 | 28.62 | 27.01 | 28.20 | 29.28 | 30.05 | 33.19 | 29.30 |
| 10 | FS10 | Frozen | 24.70 | 24.70 | 23.81 | 26.05 | 27.32 | 28.21 | 23.92 | 22.69 | 23.76 | 23.74 | 26.04 | 29.03 | 28.33 |
| 11 | FS11 | Frozen | 28.20 | 28.90 | 27.71 | 29.75 | 33.98 | 32.52 | 30.49 | 27.84 | 26.75 | 27.16 | 30.02 | 34.10 | 25.98 |
| 12 | FS12 | Frozen | 28.10 | 28.80 | 27.50 | 30.01 | 29.82 | 31.46 | 27.13 | 25.62 | 26.90 | 27.27 | 29.39 | 32.58 | 28.38 |
| 13 | FS13 | Frozen | 25.92 | 26.61 | 25.10 | 27.35 | 30.18 | 29.99 | 26.31 | 25.18 | 24.54 | 25.36 | 27.23 | 30.90 | 27.41 |
| 14 | FS14 | Frozen | 30.80 | 30.26 | 29.77 | 30.47 | 32.59 | 34.37 | 31.12 | 29.06 | 29.43 | 30.29 | 32.15 | 35.50 | 31.28 |
| 15 | FS15 | Frozen | 32.21 | 32.66 | 31.76 | 33.34 | 36.36 | 36.77 | 32.84 | 33.39 | 25.52 | 25.63 | 33.87 | 38.74 | 31.07 |
| 16 | FS16 | Frozen | 23.40 | 23.60 | 23.50 | 24.12 | 24.29 | 26.30 | 21.79 | 20.43 | 22.39 | 23.60 | 24.79 | 27.02 | 32.57 |
| 17 | FS17 | Frozen | 22.64 | 23.27 | 21.89 | 23.49 | 27.90 | 27.08 | 24.24 | 22.22 | 20.42 | 20.92 | 24.61 | 27.29 | 26.89 |
| 18 | FS18 | Frozen | 21.16 | 20.67 | 21.47 | 23.92 | 26.34 | 25.31 | 21.17 | 19.68 | 18.56 | 19.40 | 23.51 | 25.81 | 26.93 |
| 19 | FS20 | Frozen | 27.90 | 27.80 | 27.17 | 29.33 | 30.17 | 30.83 | 27.79 | 26.28 | 28.51 | 27.87 | 28.30 | 31.73 | 27.00 |
| 1 | FRS 1 | Pros pective* | 26.10 | 26.40 | 25.88 | 28.21 | 29.34 | 30.57 | 25.83 | 26.56 | 25.82 | 26.43 | 27.64 | 30.81 | 27.69 |
| 2 | FRS 2 | Pros pective* | 26.60 | 27.50 | 26.23 | 28.31 | 32.23 | 31.73 | 28.94 | 27.11 | 26.66 | 26.77 | 30.55 | 33.54 | 29.46 |
| 3 | FRS 3 | Pros pective* | 30.90 | 30.90 | 30.31 | 32.24 | 31.32 | 32.60 | 29.98 | 28.76 | 30.39 | 30.59 | 31.00 | 33.47 | 32.61 |
| 4 | FRS 4 | Pros pective* | 31.60 | 31.70 | 30.56 | 32.18 | 33.95 | 34.93 | 34.13 | 31.98 | 30.67 | 30.91 | 33.22 | 36.66 | 29.81 |
| 5 | FRS 5 | Pros pective* | 25.80 | 25.80 | 25.29 | 27.02 | 31.63 | 30.22 | 27.86 | 25.99 | 21.74 | 25.65 | 29.11 | 32.25 | 28.80 |
| 6 | FRS 6 | Pros pective* | 24.50 | 24.50 | 23.89 | 25.79 | 26.22 | 26.95 | 24.59 | 22.90 | 24.15 | 24.00 | 25.60 | 28.25 | 30.37 |
| 7 | FRS 7 | Pros pective* | 25.70 | 25.90 | 24.97 | 27.19 | 30.84 | 30.24 | 29.84 | 27.55 | 25.71 | 26.10 | 28.01 | 31.67 | 27.14 |
| 8 | FRS 8 | Pros pective* | 26.20 | 26.90 | 25.83 | 27.96 | 28.47 | 30.38 | 28.06 | 26.74 | 25.47 | 26.80 | 28.19 | 31.11 | 34.69 |
| 9 | FRS 9 | Pros pective* | 19.40 | 19.40 | 18.44 | 20.62 | 20.11 | 21.67 | 17.94 | 17.18 | 19.24 | 18.97 | 19.90 | 22.16 | 29.51 |
| 10 | FRS 10 | Pros pective* | 30.70 | 31.60 | 30.38 | 33.81 | 34.03 | 36.44 | 32.64 | 30.96 | 31.71 | 34.65 | 34.56 | 41.0 | 27.00 |
| 11 | FRS 11 | Pros pective* | 34.40 | 34.70 | 33.30 | 35.40 | 36.59 | 37.21 | 37.58 | 35.40 | 35.86 | 32.13 | 37.04 | 39.64 | 26.56 |
| 12 | FRS 12 | Pros pective* | 31.60 | 32.10 | 31.23 | 33.24 | 36.65 | 36.36 | 37.16 | 33.50 | 31.94 | 30.91 | 33.52 | 37.07 | 25.80 |
| 13 | FRS 13 | Pros pective* | 30.80 | 31.00 | 31.45 | 32.82 | 36.11 | 35.50 | 32.40 | 31.13 | 31.55 | 30.36 | 32.62 | 35.99 | 25.96 |
| 14 | FRS 14 | Pros pective* | 31.30 | 31.60 | 30.87 | 32.51 | 37.38 | 36.06 | 36.30 | 32.01 | 30.45 | 31.43 | 33.63 | 38.81 | 27.80 |
| 15 | FRS 15 | Pros pective* | 33.49 | 34.21 | 32.72 | 35.41 | 41.0 | 41.0 | 32.13 | 30.41 | 33.08 | 33.58 | 34.03 | 37.49 | 27.60 |
| 16 | FRS 16 | Pros pective* | 33.61 | 32.67 | 35.18 | 38.46 | 38.7 | 37.78 | 35.54 | 34.58 | 36.86 | 38.09 | 36.49 | 41.0 | 27.85 |
| 17 | FRS 17 | Pros pective* | 33.33 | 33.33 | 32.13 | 34.40 | 39.8 | 37.16 | 35.7 | 31.47 | 31.58 | 31.89 | 32.86 | 35.12 | 26.99 |
| 18 | FRS 18 | Pros pective* | 35.11 | 36.05 | 34.28 | 35.87 | 41.0 | 38.99 | 33.29 | 31.4 | 34.26 | 33.75 | 34.20 | 41.0 | 29.17 |
| 19 | FRS 19 | Pros pective* | 25.01 | 25.05 | 24.54 | 26.65 | 31.6 | 30.15 | 27.11 | 25.4 | 24.31 | 24.17 | 26.27 | 30.01 | 25.45 |
|  |  | Avg | 28.04 | 28.28 | 27.53 | 29.49 | 31.77 | 32.12 | 29.04 | 27.44 | 27.15 | 27.61 | 29.89 | 33.35 | 28.49 |

**Table. 3.** Summary of the C_T_ values obtained for each of the gene targets for the SORP (standard and single-tube Hemi-nested), & N1/N2 US-CDC assay for their respective gene targets. Average C_T_ values of 38 SARS-CoV-2 POS gargle samples tested on the N1/N2 and SORP assay using the purified RNA and extraction-free methods. ^‡^: Not tested; ΔC_T_:=C_T(Direct PCR)_-C_T(Extracted RNA)_.

| SARS-CoV-2 Assay | Gene Target | C<sub>T</sub><br>(n=38) |  | ΔC<sub>T</sub> |
| --- | --- | --- | --- | --- |
|  |  | Direct-PCR | Extracted-RNA | ΔC<sub>T</sub> = C<sub>T</sub>(Direct PCR) - C<sub>T</sub>(Extracted RNA) |
| N1/N2 US-CDC<br>Nucleocapsid Assay | N1 | 29.89 | 27.15 | +2.74 |
|  | N2 | 33.35 | 27.61 | +5.74 |
| Extracted SORP | S | 31.77 | 27.53 | +4.24 |
|  | ORF8 | 32.12 | 29.49 | +2.63 |
| STHN SORP | S | 29.04 | Not Tested<sup>†</sup> | +1.51 |
|  | ORF8 | 27.44 | Not Tested<sup>†</sup> | -2.05 |

**Fig. 2.**
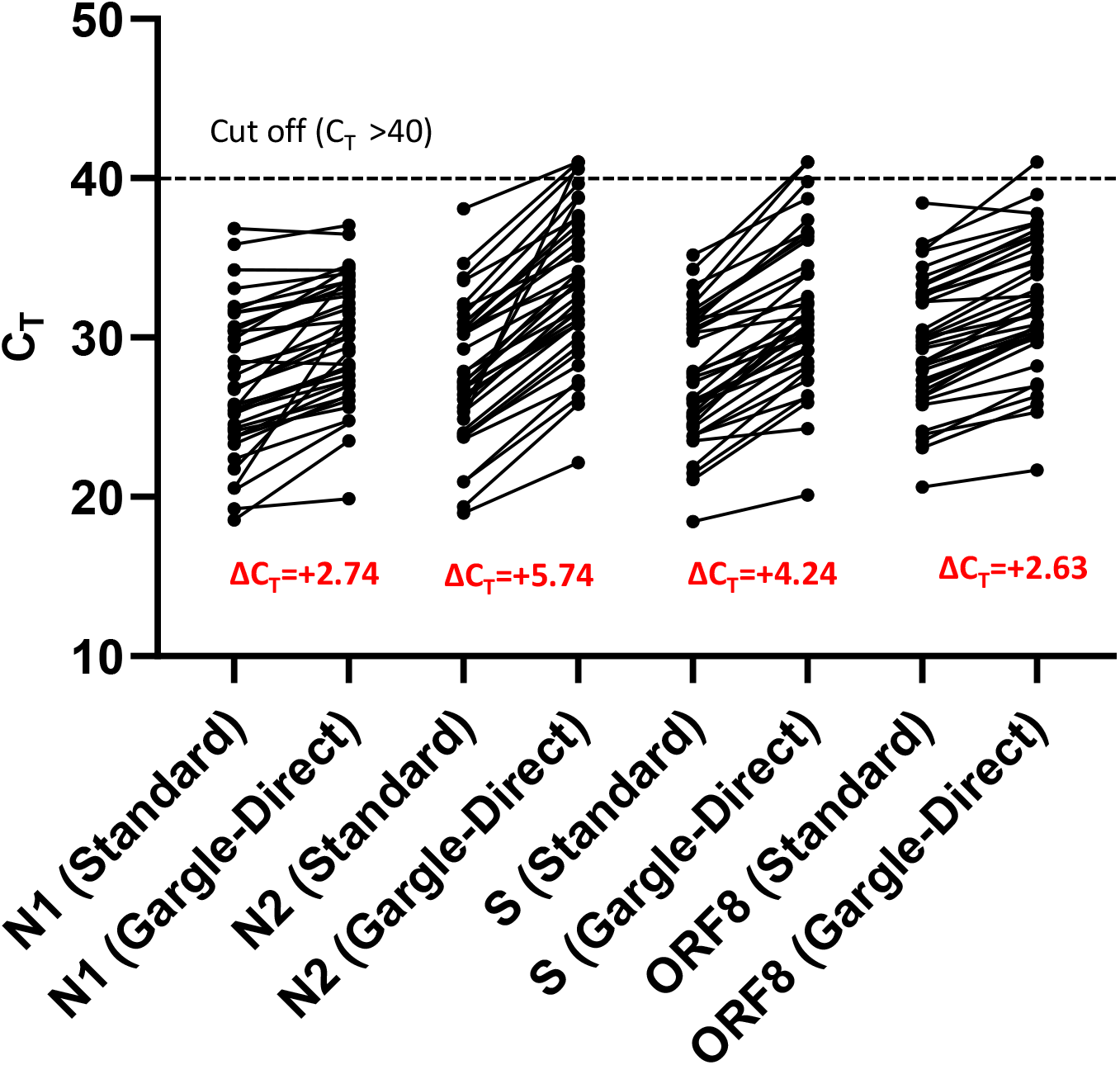
Diagram depicting the paired C_T_ values obtained by the standard RNA extraction-based and Gargle-Direct processes for the N1, N2, S and ORF8 gene targets. Values over the cut-off line represent undetected values and have been assigned a C_T_ value of 41.0. The dotted line represents the C_T_ cut-off for positivity. ΔC_T_:=C_T(Direct PCR)_-C_T(Extracted RNA)_.

When these 38 positive saline mouth rinse/gargle samples were tested on the SORP assay using the standard RNA extracted protocol, both S and ORF8 gene targets were detected in all the samples, including the RNase P internal control (Table. 2). The average C_T_ values across all the 38 positives saline gargle samples using the standard RNA extraction-based method were: S=27.53 and ORF8=29.49. For the same cohort of positive gargle samples tested on the Gargle-Direct approach, both the S and ORF8 were detected in all samples, except in one sample FRS15 (S/ORF=NEG) which was negative for both the targets. One sample, FRS18 was positive for only a single target (S=NEG/ORF=POS) (Table. 2), making it indeterminate. The average C_T_ values for the S and ORF8 gene targets were: 31.77 and 32.12 respectively (Table. 2). This represented a net C_T_ increase for the S (ΔC_T_= +4.24) and ORF8 (ΔC_T_=+2.63) gene targets, when compared between the extraction-free and the standard RNA extracted approach (Table. 3). This difference was statistically significant using two-sided paired-sample t test (P<0.0001) for both these gene targets (Fig. 2). RNAse P was consistently detected in all the positive 38 gargle samples tested using the extraction-free protocol. No false positives were detected amongst the 75 SARS-CoV-2 negative mouth rinse/gargle samples tested with the Gargle-Direct protocol. Overall, the performance of the SORP triplex assay on the Gargle-Direct protocol was: sensitivity of 97.37% (95%CI:86.19% to 99.93%), specificity of 100% (95%CI:95.20% to 100.0%), Accuracy: 99.12% (95%CI:95.17% to 99.98%).

When the 38 SARS-CoV positive mouth rinse/gargle samples were tested on the Gargle-Direct protocol using the STHN-RT-qPCR SORP assay, the average C_T_ values recorded for the S and ORF8 gene targets were, 29.04 and 27.44 respectively (Table. 2). This represented a ΔC_T_ of +1.51 and −2.05 for the S and ORF8 genes respectively, when compared with the standard RNA extracted method (Fig. 3). There were no false positives detected amongst the 75 SARS-CoV-2 negative mouth rinse/gargle samples tested on the STHN-RT-qPCR SORP assay. RNase P was detected in all the 38 positive and 75 negative samples tested on the Gargle-Direct protocol using the STHN-RT-qPCR. Overall, the SORP STHN-RT-qPCR triplex assay displayed 100% concordance with the BCCDC-Ref assay and similar sensitivity/specificity with the N1/N2 US-CDC assay when used in standard approach (extracted RNA) (Table 2.).

**Fig. 3.**
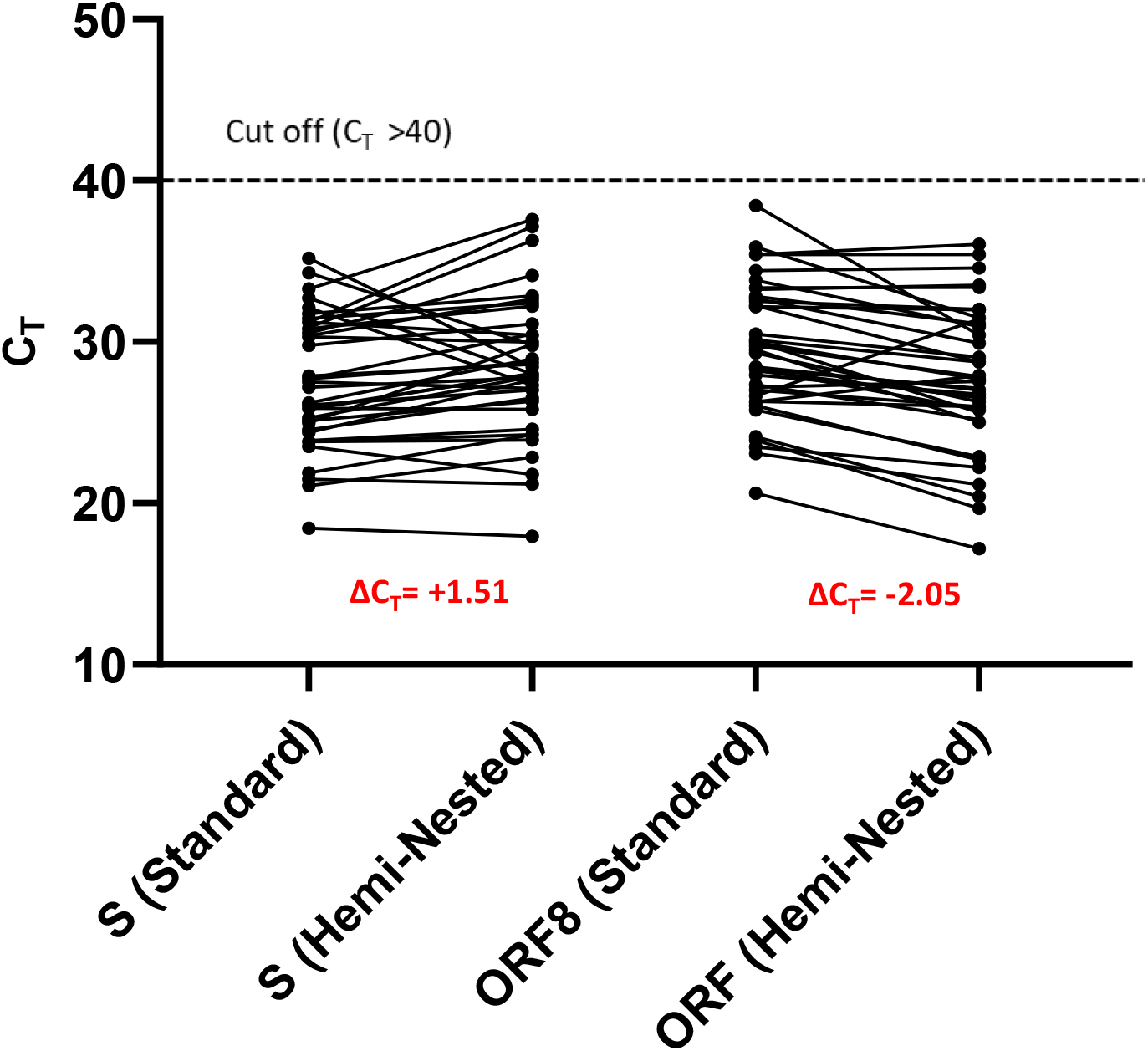

## DISCUSSION

In our study, we evaluated a convenience sample of 38 SARS-CoV-2 positive and 75 SARS-CoV-2 negative saline mouth rinse/gargle patient specimens. The positive samples represent a range of viral loads, with approximately 40% having relatively low amounts of SARS-CoV-2 RNA (C_T_> 30). The standard assays used to test these and other sample types are based on an RNA extraction-based approach, which presents multiple challenges for high-throughput processing. To address the bottlenecks in implementing mass testing using this standard approach, we decided to explore the possibility of using an “extraction-free” approach, whereby the saline mouth rinse/gargle sample could be directly added to the RT-qPCR reaction mixture with minimal pre-analytic handling.

While developing the “extraction-free” approach, we found two main attributes of the saline mouth rinse/gargle sample type that helped us to reduce the pre-analytical steps. First, in terms of sample complexity, a saline mouth rinse/gargle sample, unlike saliva, is a dilute, homogenous sample, with a water-like consistency. This makes it more amenable to liquid handling - both manual and automated. No pre-dilution, or addition of any liquefaction agents, was necessary, allowing mouth rinse/ gargle sample to be directly added into the RT-qPCR reaction. Secondly, total ionic strength of sodium chloride (0.154 M) in the saline solution was found to be compatible with the amplification polymerase, especially the reverse transcriptase enzyme that is known to be sensitive to inhibitors, used in our Gargle-Direct protocol. Based on preliminary study, we found that amongst the commonly available master mixes used for SARS-CoV-2 RT-qPCR, the Luna™ One-Step Universal RT-qPCR master mix, performed better in presence of potential inhibitor(s) containing templates, than some of the other RT-qPCR mixes for e.g. the 4X TaqMan Fast Virus 1-step Master Mix. This observation is consistent with other studies where Luna™ One-Step Universal RT-qPCR master mix, successfully amplified SARS-CoV-2 from unpurified templates, prepared from sample types recommended for COVID-19 testing for e.g. NPS (Bruce et al., 2020), saliva (Vogel et al., 2020) and mouth washes (Maricic et al., 2020).

The compatibility of the saline gargle sample type for direct amplification helped us to eliminate the need for a preliminary dilution step, as is commonly required for viral transport media (VTM) based sample types, to make the sample compatible with the RT-qPCR amplification chemistry (Hasan et al., 2020). The only pre-analytical step we chose to perform was a simple heat inactivation step (65 °C for 20 minutes) to inactivate the SARS-CoV-2 virus. This was done so that the sample would be safe for downstream robotic pipetting during the RT-qPCR reaction. The choice of a relatively low inactivation temperature was chosen in our workflow as high temperature of lysis (90 to 95 °C) has been shown to be detrimental to the stability of the SARS-CoV-2 RNA (Pastorino et al., 2020). Direct addition of the sample without heat inactivation was also tested and gave comparable results (data not shown). To address the issue of a decline in sensitivity due to the direct addition of a crude amplification template, we developed a hemi-nested version of the SORP RT-qPCR assay. To prevent contamination and increase the testing throughput, the hemi-nested RT-qPCR was performed in a “single-tube” configuration, where both the primary and secondary PCR cycling were carried out in the same reaction well without opening the plate/well. This not only reduced handling steps but also alleviated any possibility of amplicon contamination in the laboratory. This two step PCR cycling process, resulted in the detection of the target with a reduced ΔC_T_ difference, for both the S and ORF8 gene targets, when tested using this novel RT-qPCR system. For the two-step cycling to work, we had to raise the melting temperature (Tm), of the pre-amplification primers (Spike-F1-LNA/Spike-Rout-2-LNA/ORF-8-F1-LNA/ORF-8-Rout-1-LNA) used in primary PCR; Table. 1). This was achieved by incorporating 2-3 LNA’s in the oligonucleotide backbone, resulting in an increase in Tm of 4-8 °C (Obika et al., 1998; Singh et al., 1998). As a result of the raised Tm, the pre-amplification (15 cycles), could be carried out at higher (70 °C) annealing temperatures. After the pre-amplification step, the PCR cycling was brought to a lower annealing temperature (60 °C) which, was the optimized temperature for the standard TaqMan™ forward/reverse primer and probes.

This study is the first to describe extraction-free RT-qPCR for diagnosis of SARS-CoV-2 using saline mouth rinse/gargle specimens, which is a promising approach for scaling up testing during the COVID-19 pandemic. Our evaluation includes the assessment of performance across multiple assays, including the FDA-authorized US-CDC assay. Limitations of this study include the relatively small number of positive specimens although, we have included more than the 30 positive and negative clinical samples recommended by the FDA for evaluation of new molecular assays as part of the Emergency Use Authorization authority. Future studies should include larger numbers of prospectively tested specimens, especially those with low viral loads (C_T_>35), to confirm these findings and assess the operational feasibility of implementing larger scale robotic Gargle-Direct testing of saline mouth rinse/gargle specimens. For example, adjusting the protocol such that heat inactivation occurs in the primary collection tube, would likely be feasible and significantly simplify the workflow.

In summary, we describe a sensitive and specific diagnostic method for SARS-CoV-2 that is operationally simple, bypasses multiple supply chain bottlenecks, utilizes a self-collected swab independent sample type, is appropriate for large scale testing, is cost effective, and can be readily adopted by other laboratories. This STHNRT-qPCR could also be applied in other SARS-CoV-2 testing scenarios where a loss of sensitivity is routinely expected e.g. SARS-CoV-2 testing by sample pooling. This would facilitate large scale SARS-CoV-2 testing, a crucial tool for the control the COVID-19 pandemic.

## Data Availability

The authors confirm that the data supporting the finding are available in the article and/or its supplementary material

## FUNDING SUPPORT

The study was supported by the Peter Wall Institute of Advanced Studies, Vancouver and the Provincial Health Services Authority (PHSA) of British Columbia.

## CONFLICT OF INTEREST STATEMENT

The authors declare no conflict of interest.

## Notes

### Competing Interest Statement

The authors have declared no competing interest.

### Author Declarations

This was approved by the BC Children's and Women's Research Ethics Board (H20-02538).

